# The Impact of COVID-19 Pandemic on The Preventive Services in Qatar

**DOI:** 10.1101/2020.09.08.20189555

**Authors:** Mohamed Ghaith Al-Kuwari, Mariam Ali Abdulmalik, Hamad Rashid Al-Mudahka, Ahmad Haj Bakri, Wadha Ahmed Al-Baker, Shaikha Sami Abushaikha, Mujeeb Chettiyam Kandy, John Gibb

## Abstract

**Background:** In March 2020, Qatar started reporting increased numbers of COVID-19 cases. At that stage, containment measures were put in place. The health authority in Qatar developed an emergency action plan to respond to the outbreak with the Primary Health Care as the main component of that response and suspended all non-urgent services including preventive health services. The aim of the retrospective analysis to measure the Impact of COVID 19 on the preventive services provided in Qatar.

**Methods:** A retrospective data analysis was conducted for all the preventive services utilization volume across the 27 PHCC health centers from the 1st of January 2017 to the 31st of July 2020.

**Results:** With 17,012 no-show appointments, well-baby and Immunization services utilization demonstrated a reduction of 40% in May and started to come back to volumes higher than expected in June. The number of cancelled appointments for breast cancer and colorectal cancer screening programs were 3,481 and 5,854 respectively. The expected volumes demand has dropped by 100% in comparison to 2017 demand. Wellness services only met 20% of its projected utilization in April, however, the services picked up in June.

**Conclusion:** These findings will guide the public health policymakers to understand the effects COVID-19 on preventive services and the risk of having an increased number of outbreaks for childhood communicable disease, cancer cases with delayed diagnosis due to the screening services suspension. In addition, the plan will address the increased number of sedately behavior due to the service’s reduced utilization of wellness services.

## Introduction

The international community is actively mobilizing all its efforts to limit the spread of severe acute respiratory syndrome coronavirus 2 (COVID-19) and reduce mortality. As of 17^th^ of August 2020, approximately 21.2 million cases and over 761,000 deaths have been reported globally and estimates of the death numbers in the future in the millions^1,2^

Governments are continuously responding at national, regional, and global levels, and health guidance for health systems and the public are being developed^3^. In assessing their options, policymakers must consider not only the immediate health effects of the pandemic but also the indirect effects of the pandemic and the response to it. An analysis of the Ebola outbreak in West Africa in 2014 demonstrated that the indirect effects of the outbreak were more severe than the outbreak itself^4^. Although mortality rates for COVID-19 seem to be low in women of reproductive age and in children^5^, these groups might be affected by the disruption of the routine health services disproportionately.

In past epidemics, health systems have struggled to sustain the routine services and utilization of services has dropped^6^. As WHO indicates, people, efforts, and medical supplies all shift to respond to the emergency. This often leads to the neglect of basic and regular essential health services. People with health problems unrelated to the epidemic find it harder to get access to health care services^7^.

During the outbreak, health workers, equipment, and facilities have been reallocated to address the influx of patients with COVID-19^8^. Restructuring of the health system could result in the closure of some health facilities, as seen in the 2014 Ebola virus outbreak^9^. The workforce in health has been additionally reduced by nosocomial COVID-19 infection and burnout^10^. Government restrictions on movement and halting non-essential activities and enforcing stay home policies might add barriers to health care accessibility^11^. Demand for preventive services might decline as concerns over COVID-19 transmission alter the perceived risk-benefit calculation for persons deciding to seek care^12^

Before COVID-19, Qatar had invested in preventive health services provided through the Primary Health Care Corporation (PHCC) to enhance healthier lives for its target population with wide health promotion and preventive services provided with the focus put on screening, healthy lifestyle promotion, and immunization.

Since March 2020 when Qatar started reporting an increasing number of COVID-19 cases with a total of 116,224 cases and 193 deaths have been reported as of August 20, 2020^13^ At the early phase, national-level preventive measures were put in place to decrease the level of transmission. To decrease the risk of infection to the patients and health care workers, Primary Health Care in Qatar had to cancel appointments for some services. This cancellation included most of the preventive health services e.g. cancer screening and wellness, while PHCC has continued the services provision of the Well Baby clinic and immunization for all under-5 years old children in Qatar.

To maintain the accessibility to the health service, primary health care centers have initiated telemedicine consultations for most of the clinical services to support the adherence to the recommendations to stay at home to minimize the risk of infection. Smoking cessation counseling was one of these services that use telemedicine consultations to compensate for the canceled appointments. Therefore, the aim of this review is to measure the Impact of COVID 19 on preventative services provided in Qatar.

## Methodology

A retrospective data analysis was conducted for all the preventative services utilizations volume across the 27 PHCC health centers as part of the practical impact analysis reporting done to monitor the service utilization during the pandemic. The data was extracted daily for the records to the statistical reporting system of health intelligence, which includes all the appointments and walk-in visits for all health centers.

The volumes of visits were categorized per the type of preventative service and in defined duration as following: visits between 1st of January 2017 and 31st of July 2020 for the cancer screening programs, and this includes breast and colorectal cancers. For wellness services, which include smoking cessation and exercises therapy, data was extracted between the 1st of January 2018 and 31st of July 2020. Well-baby services comprise immunization, growth development monitoring, and other screening e.g. hearing and hip dysplasia. The wellness data was extracted between the 1st of January 2018 and 31st of July 2020.

The number of missed appointments were calculated from the cancelled appointments for the cancer screening and wellness. For the well-baby clinic, number of no-show appointments were considered as missed appointments. The utilization trends were established to track the differences before and during the COVID-19pandemic in 2020 in compression to the previous years and the projected trend calculated for 2020.

## Results

From mid-March till July 2020, health centers canceled all cancer screening and wellness appointments, while well-baby clinic was the only preventive service that was carried out during the pandemic. Table-1 shows that the number of no-show appointments for Well-baby clinic was 17,012. The number of cancelled appointments was more in colorectal cancer screening than breast cancer screening, 5,854 vs, 3,481. While the number of canceled wellness appointments was 3,385.

**Table-1.** Number of missed appointments per type of the preventive service March-July 2019.

| Preventive service | Missed appointments |
| --- | --- |
| Breast cancer screening * | 3,481 |
| Colorectal cancer screening * | 5,854 |
| Well-baby & immunization clinic** | 17,012 |
| Wellness * | 3,385 |

\*Cancelled appointments.
\*\*No-show appointments.

The breast cancer screening program has around a length of 3 years, so expected volumes will correlate with the screening demand from 2017 and since April 2020 the service utilization has dropped by 100% with the loss of 5,100 individuals to be screened as shown in figure-1.

**Figure-1:**
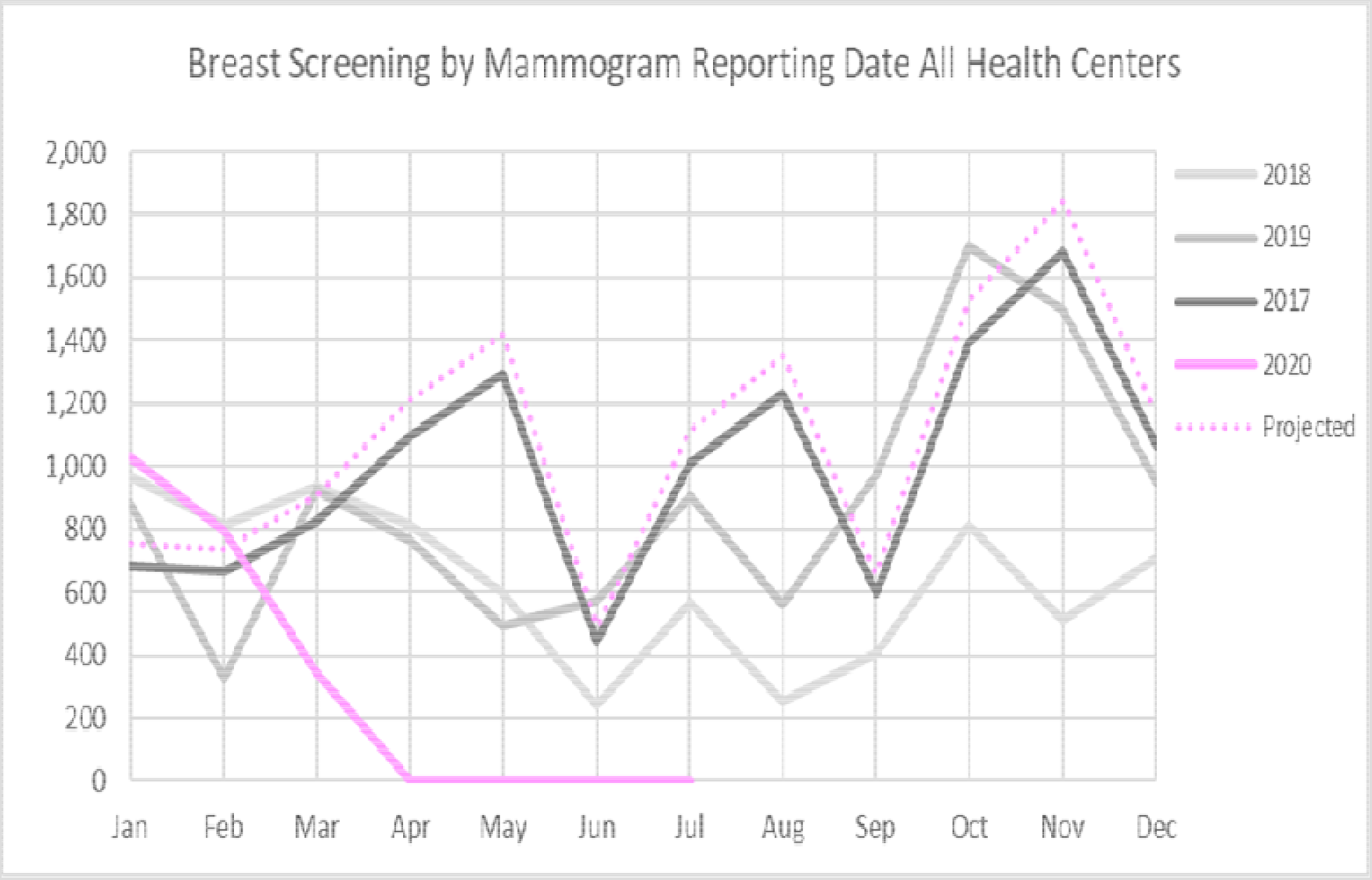
Breast cancer screening services across all health centers between January 2017 and July 2020

Colorectal cancer screening services has a round of one year between 2016 and 2018 then the round was changed to two years as of 2019, the services were suspended during the COVID-19 pandemic as of Mid-March 2020, the latter resulted in a drop of the service utilization by 100% from April to July 2020 in comparison to other years as demonstrated in figure-2.

**Figure-2:**
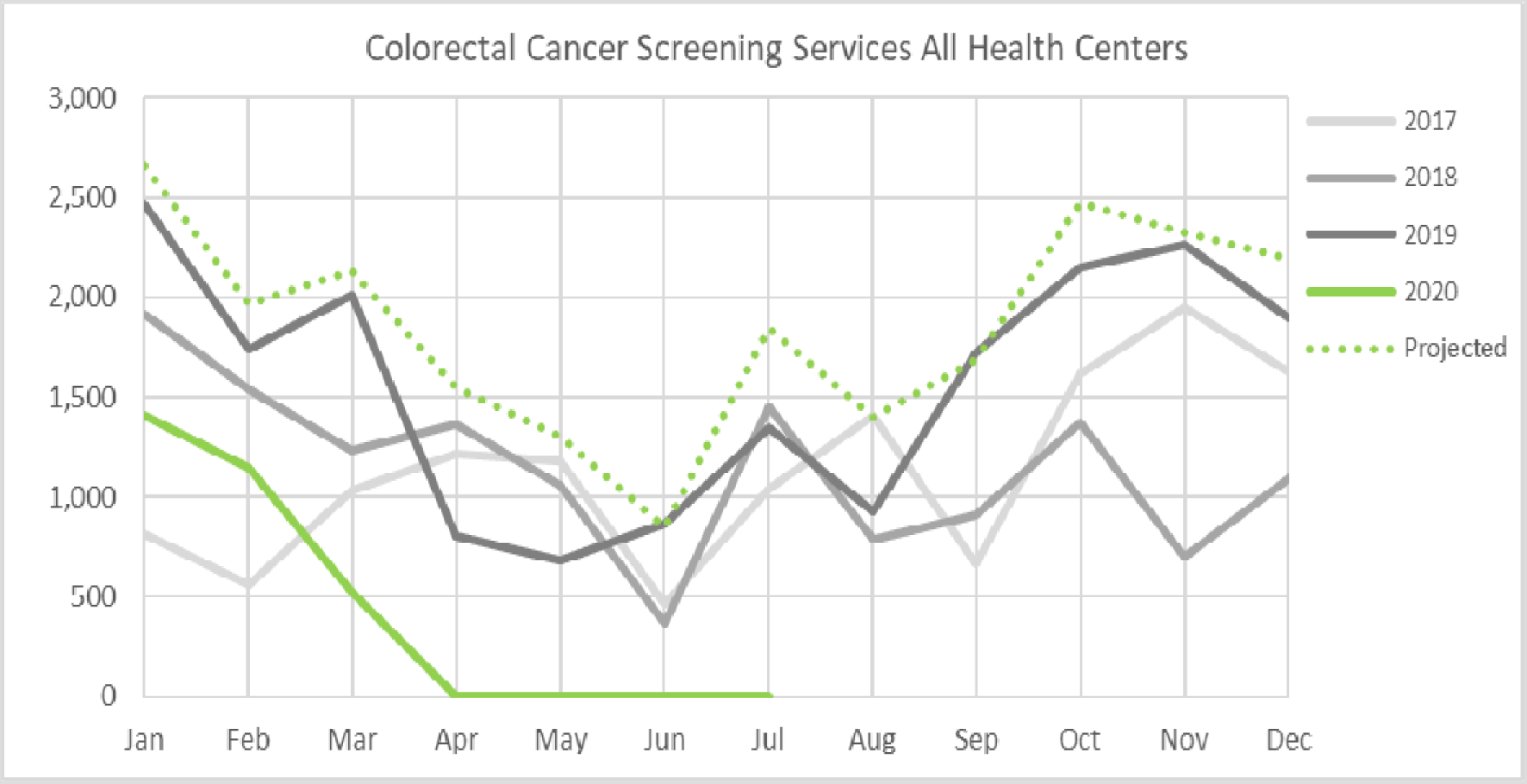
Colorectal cancer screening services across all health centers between January 2017 and July 2020

Figure-3 shows that the Well-baby and immunization services demonstrated a decline in the utilization of the services between March and May 2020 to almost 40% reduction in May 2020 in comparison to the same month in 2019. However, as of the beginning of June 2020, appointments have come back to volumes higher than expected yet to decline slightly towards the end of July due to the Eid’s public holiday.

**Figure-3:**
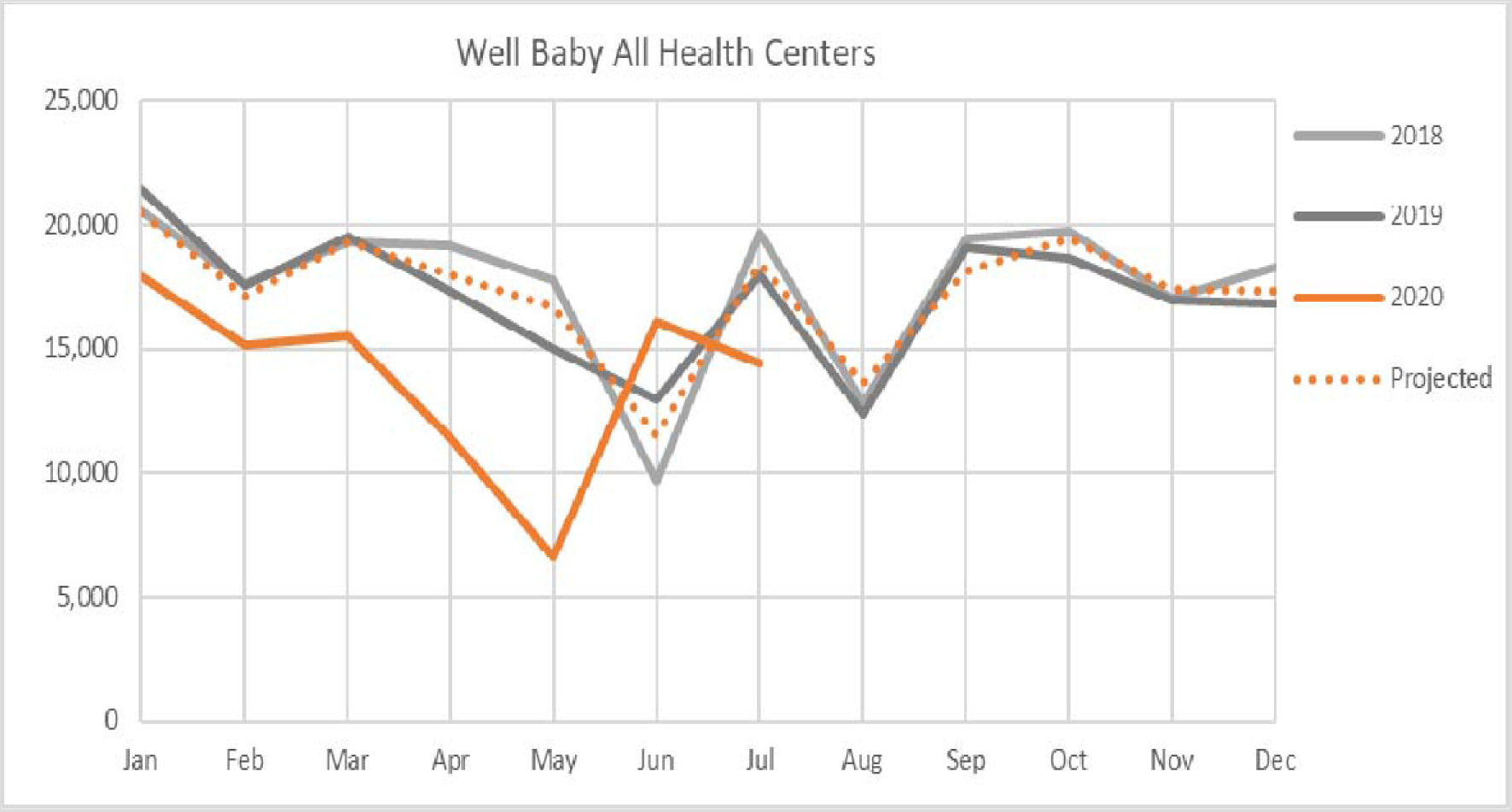
Well baby services utilization across PHCC health centers between January 2018 and July 2020.

Wellness services demonstrated a steep decline in the utilization of the services between March and May 2020 to almost 20% of the expected utilization for the month of April as shown in figure-4. However, there was a steady increase in services utilization during the month of June 2020. The latter might be attributed to the increased utilization of the wellness services teleconsultation, mainly smoking cessation virtual services as shown in figure-5. However, there was a slight incline in the tobacco cessation virtual services in the month of July due to public holidays.

**Figure-4:**
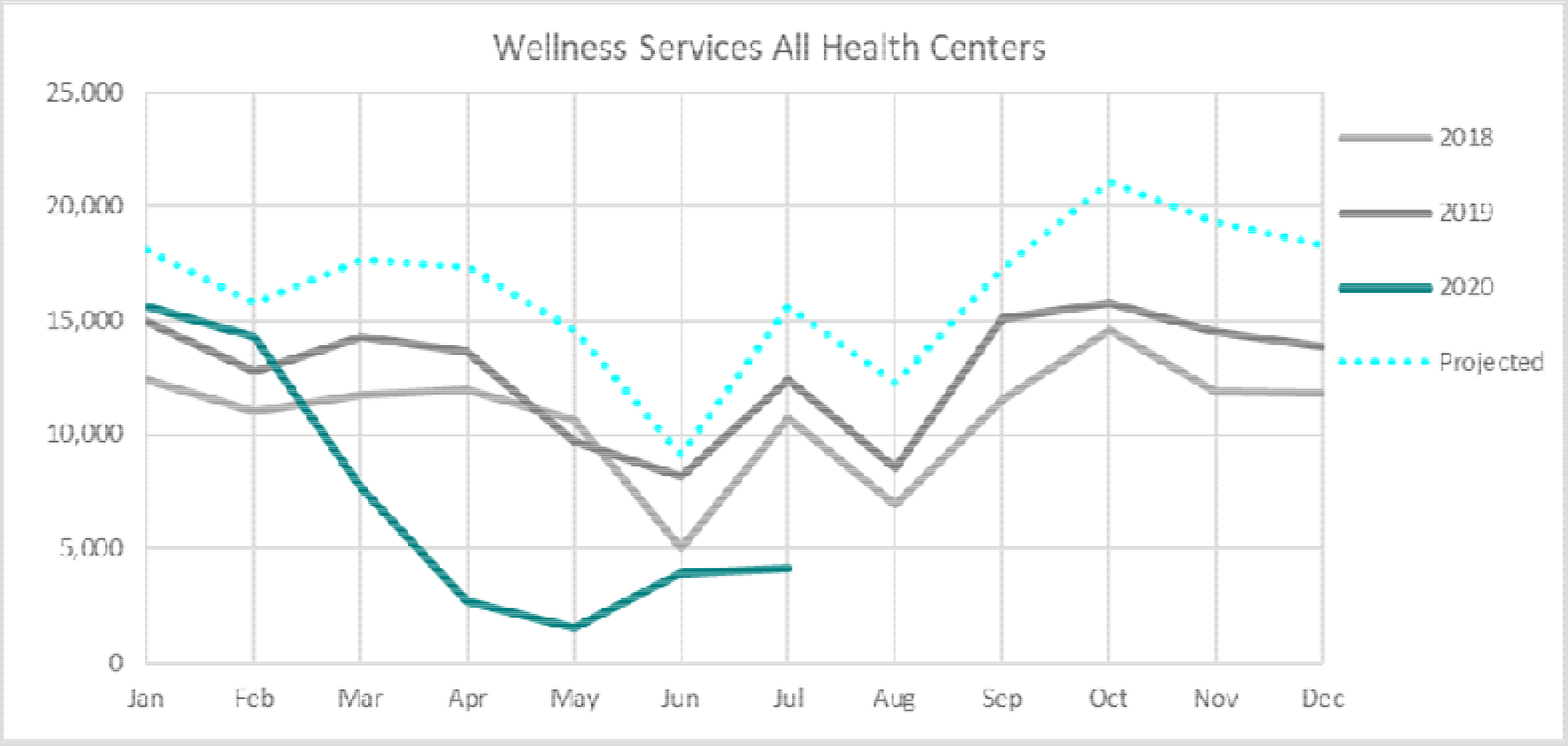
Wellness services utilization across PHCC health centers between January 2018 and July 2020

**Figure-5:**
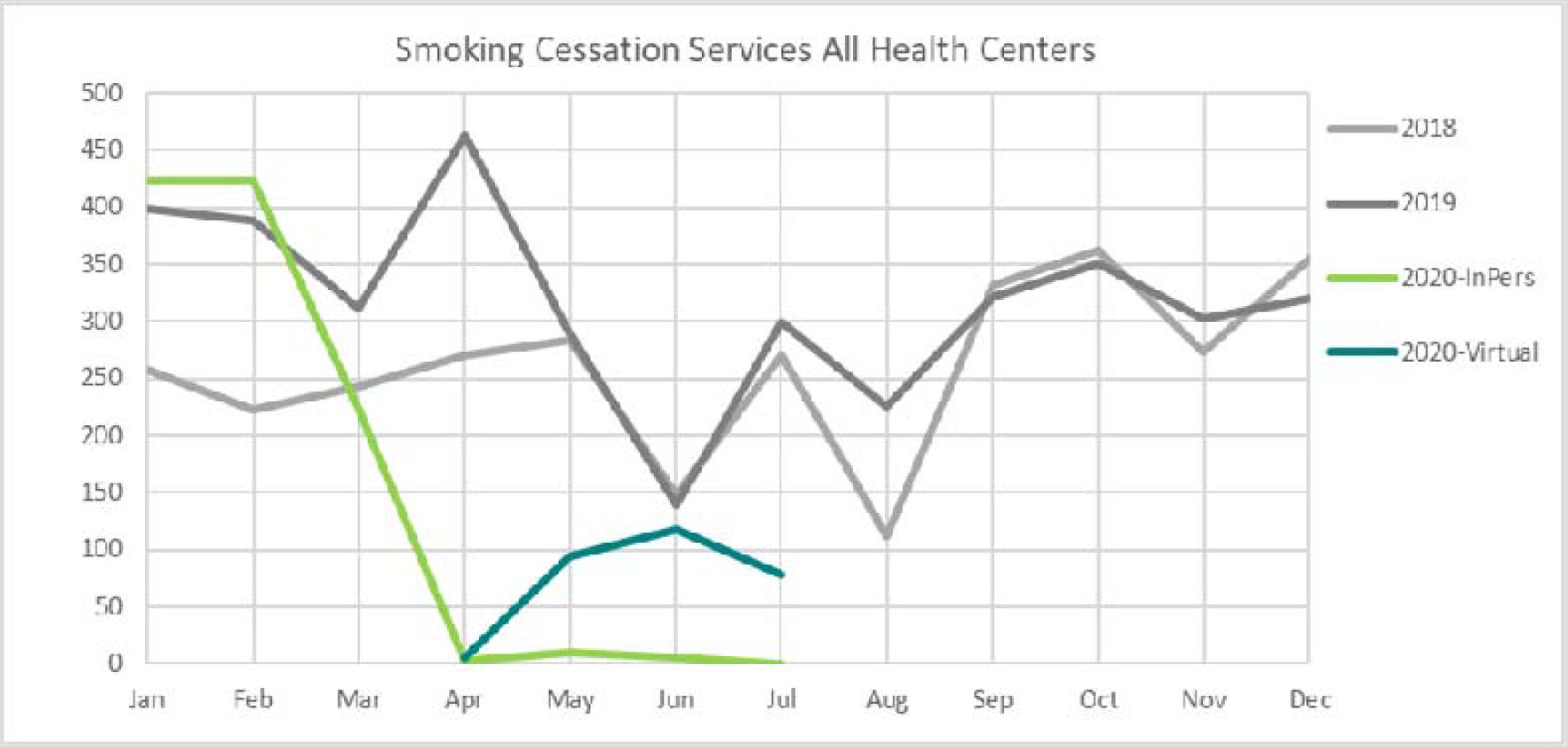
Tobacco cessation services utilization across PHCC health centers between January 2018 and July 2020

## Discussion

The COVID-19 pandemic response has substantial consequences on health services in Qatar. most non-urgent appointments and non-urgent services were suspended to decrease infection to the patients, which had major impact on the utilization of the preventive services.

The cancer screening program for breast and colorectal cancer has been suspended as of 11th of March 2020 with a 100% drop in the utilization of the services with more than 9,000 appointments for screening were cancelled between April and July 2020. A similar pattern was reported in Italy and the UK, due to national social distancing policy and allocating all health care professionals and the other resources to manage COVID-19 cases^14,15^.

The effect of this pause in the cancer screening might be more noticeable after extended periods of follow up, when we see a delay in cancer diagnosis presented in form of having more cases diagnosed in the late stage and reduction of the cases diagnosed in the early stage in comparison to the previous years. The long-term effect, which includes mortality indicators might need 5 years to be detected^16^.

Well-baby and immunization service were the only preventive service that provided as it was before the pandemic. However, it showed a sharp decline in the utilization of the services between March and May 2020 to almost 40% reduction in May 2020 in comparison to the same month in 2019 with more than 17,000 children did not show for their appointments. The decline in immunization uptake has been reported in different countries during the pandemic^17^. Similar phenomena have been reported after the Ebola virus outbreak followed by measles outbreaks among children in Africa^18^. This might be attributed to the fear factor of exposing children to the outside environment and keeping them safe at home during the COVID-19 pandemic.

Moreover, the well-baby clinic provides good screening opportunities at the national level to detect a number of physical and devolvement conditions among infants and preschool children e.g. growth and developmental delay, hip dysplasia, and deafness. Missing visits during the COVI-19 pandemic without may result in missing theses these cases that need early management. Nevertheless, since the 4th of June 2020 when the health authority announced the phasing stages of lifting the restrictions imposed due to the COVID-19 pandemic, the utilization volume for well-baby services started to pick up as a result of using the recall system for the defaulters.

The wellness services uptake has been affected by the COVID-10 pandemic since there was a steep decline in the utilization of the services between March and May2020. However, with the introduction of virtual consultation as of April 2020 for the smoking cessation services only, the wellness services started to pick up again. The other services for lifestyle intervention such as nutrition and exercise counseling remained suspended. The expected long-term effect on cardiometabolic markers would an area for investigation, especially that the wellness centers stopped the gym and swimming pools used for exercise therapy for patients with chronic conditions e.g. diabetes, hypertension, obesity, and arthritis. This long interruption on the exercise classes will produce a negative impact on the clinical outcome and the quality of life of the patients.

These findings will guide the public health policymakers to understand the effects COVID-19 on preventive services through monitoring the utilization of the essential preventive health services during any pandemic and putting the recovery plans to manage the backlog in the appointments. This recovery plan should take into consideration the assessment of the patients with chronic conditions and their physical activity and smoking status to access the wellness service. An extensive plan to increase the capacity of the service to screen the largest number of individuals who missed the cancer screening should be a priority in that plan. Measuring the risk of having an increased number of cancer cases with delayed diagnosis due to the screening services suspension needs to be considered in the outcome of the screening program. In addition, the CDC surveillance system should monitor the increased number of immunization program defaulters due to the services reduced utilization and follow up the effect of the recall system by geography to identify the possible areas that need more intensive catch up immunization campaigns.

## Data Availability

All the data referred to in the manuscript is available.

